# Quantifying the information in noisy epidemic curves

**DOI:** 10.1101/2022.05.16.22275147

**Authors:** Kris V Parag, Christl A Donnelly, Alexander E Zarebski

## Abstract

Reliably estimating the dynamics of transmissible diseases from noisy surveillance data is an enduring problem in modern epidemiology. Key parameters, such as the instantaneous reproduction number, *R*_*t*_ at time *t*, are often inferred from incident time series, with the aim of informing policymakers on the growth rate of outbreaks or testing hypotheses about the effectiveness of public health interventions. However, the reliability of these inferences depends critically on reporting errors and latencies innate to those time series. While studies have proposed corrections for these issues, methodology for formally assessing how these sources of noise degrade *R*_*t*_ estimate quality is lacking. By adapting Fisher information and experimental design theory, we develop an analytical framework to quantify the uncertainty induced by under-reporting and delays in reporting infections. This yields a novel metric, defined by the geometric means of reporting and cumulative delay probabilities, for ranking surveillance data informativeness. We apply this metric to two primary data sources for inferring *R*_*t*_: epidemic case and death curves. We find that the assumption of death curves as more reliable, commonly made for acute infectious diseases such as COVID-19 and influenza, is not obvious and possibly untrue in many settings. Our framework clarifies and quantifies how actionable information about pathogen transmissibility is lost due to surveillance limitations.

## Introduction

The *instantaneous reproduction number*, denoted *R*_*t*_ at time *t*, is an important and popular temporal measure of the transmissibility of an unfolding infectious disease epidemic [1]. This parameter defines the average number of secondary infections generated by a primary one at *t*, providing a critical threshold for delineating growing epidemics (*R*_*t*_ *>* 1) from those likely to become controlled (*R*_*t*_ *<* 1). Estimates of *R*_*t*_ derived from surveillance data are widely used to evaluate the efficacies of interventions [2, 3] (e.g., lock-downs), forecast upcoming disease burden [4, 5] (e.g., hospitalisations), inform policymaking [1] and improve public health awareness [6].

The reliability of these estimates depends fundamentally on the quality and timeliness of the surveillance data available. Practical epidemic monitoring is subject to various errors or imperfections that can obscure or bias inferred transmission dynamics [7]. Prime among these are *under-reporting* and *reporting delays*, which can scale and smear *R*_*t*_ estimates, potentially misinforming public health authorities [8, 9]. Under-reporting causes some fraction of infections to never be reported, while delays redistribute reports of infections incorrectly across time. The ideal data source for estimating *R*_*t*_ is the time series of new or incident infections, *I*_*t*_.

Unfortunately, infections are difficult to observe directly and proxies such as reported cases, deaths, hospitalisations, prevalence and viral surveys from wastewater must be used to gauge epidemic transmissibility [1, 10]. Each of these data streams provides a noisy approximation to the unknown *I*_*t*_ but with distinct and important relative advantages. We focus on the most popular ones: the epidemic curve of reported cases, *C*_*t*_ at time *t*, and that of death counts, *D*_*t*_, and investigate how their innate noise sources differentially limit *R*_*t*_ inference quality.

The epidemic case curve, *C*_*t*_, records the most routinely available data i.e., counts of new cases [11], but is limited by delays and under-reporting. Ascertainment delays smear or reorder the case incidence and may emerge from fixed surveillance capacities, weekend effects and lags in diagnosing symptomatic patients (e.g., the time from infection to a positive test) [8, 12]. Delays may be classed as *occurred but not yet reported* (OBNR), when source times of delayed cases eventually become known (i.e., delays cause right censoring of the case counts), or what we term as *never reported* (NEVR), when source times of past cases are never uncovered [13–15].

Case under-reporting or under-ascertainment strongly distorts the true, but unknown, infection incidence curve, altering its size and shape [9, 16]. Temporal fluctuations in testing intensity, behaviour-based reporting (e.g., by severity of symptoms) [17], undetected asymptomatic carriers and other surveillance bottlenecks can cause under-ascertainment or inconsistent reporting [18, 19]. *Constant reporting* (CONR) describes when the case detection fraction or probability is stable. We term the more realistic scenario in which this probability varies appreciably with time as *variable reporting* (VARR).

Death time series, *D*_*t*_, count newly reported deaths attributable to the pathogen being studied and are also subject to under-reporting and reporting delays, but with two main differences [10]. First, death reporting delays incorporate an extra lag for the intrinsic time it takes an infection to culminate in mortality (this also subsumes hospitalisation periods). Second, apart from the under-reporting fraction of deaths, there is another scaling factor known as the infection fatality ratio, which defines the proportion of infections that result in mortality [2, 20]. We visualise how the noise types underlying case and death curves distort infection incidence in Fig. 1.

**Fig. 1:**
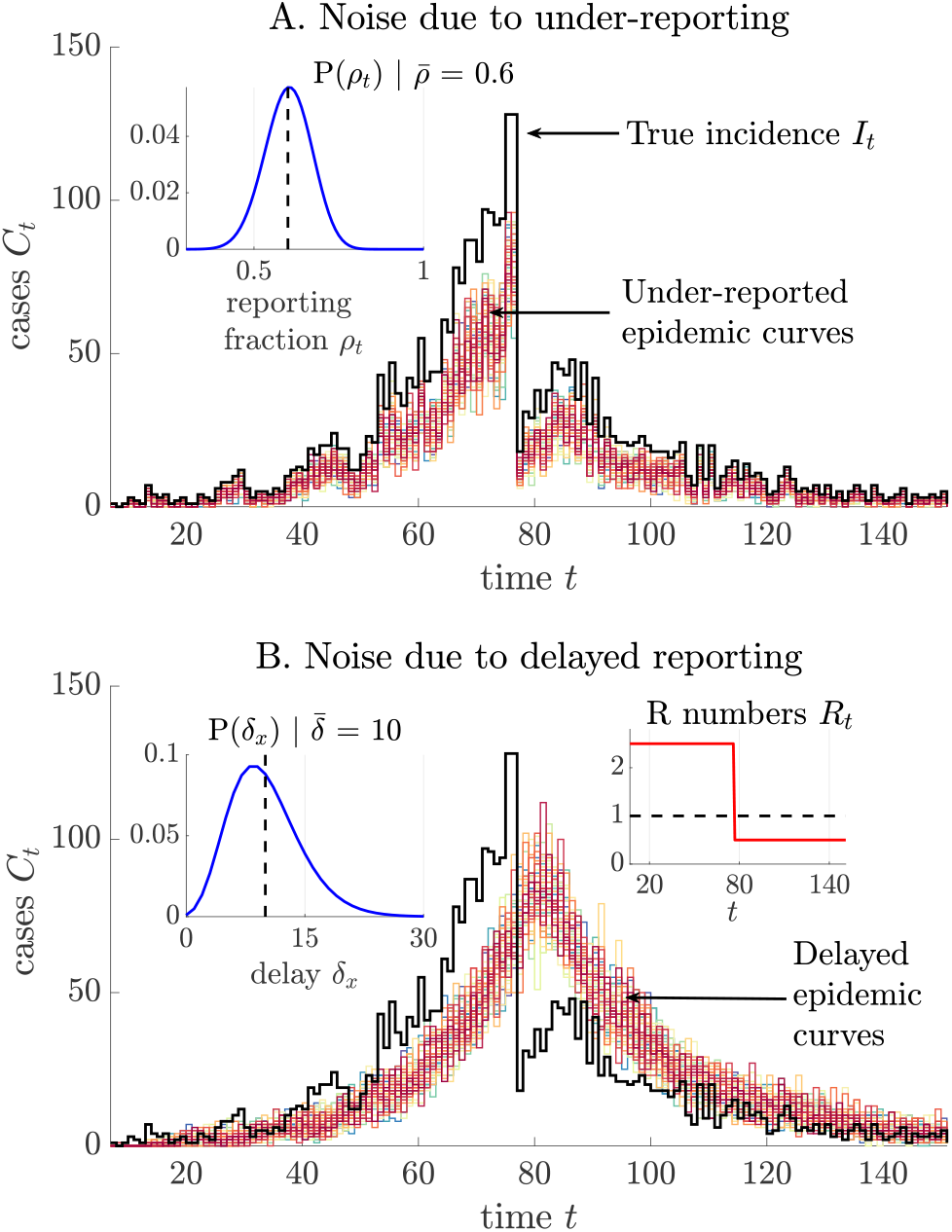
Under-reporting and delayed reporting noise. We simulate true infection incidence *I*_*t*_ (black) from a renewal model (Eq. (15) with Ebola virus dynamics) with reproduction number *R*_*t*_ that switches from supercritical to subcritical spread due to an intervention. Panel A shows under-reported case curves (50 realisations, various colours) with reporting fractions sampled from the distribution in the inset. We observe stochastic trajectories and appreciable under-counting of peak incidence. Panel B considers delays in case reports (50 realisations, various colours) from the distribution plotted in the inset. We find variability and a smearing of the sharp change in incidence due to *R*_*t*_ (also provided as an inset). The main question of this study is how do we quantify which of these two scenarios incurs a larger loss of the information originally available from *I*_*t*_, ideally without simulation.

Although the influences of surveillance latencies and under-ascertainment fractions on key parameters, such as *R*_*t*_, are known [8, 19, 21, 22] and much ongoing work attempts to compensate for these noise sources [10, 23–25], there exists no formal framework for assessing and exposing how they inherently limit information available for estimating epidemic dynamics. Most studies utilise simulation-based approaches (with some exceptions e.g., [9, 22]) to characterise surveillance defects, which while invaluable, preclude generalisable insights into how epidemic monitoring shapes parameter inference.

Here we develop one such analytic framework for quantifying the information within epidemic data. Using *Fisher information* theory we derive a measure of how much usable information an epidemic time series contains for inferring *R*_*t*_ at every time. This yields metrics for cross-comparing different types of surveillance time series as we are able to explicitly quantify how under-reporting (both CONR and VARR) and reporting delays (exactly for OBNR with a tight upper bound for NEVR) degrade available information. As this metric only depends on the properties of surveillance (and not *R*_*t*_ or *I*_*t*_) we extract simulation-agnostic insights into what are the least and most detrimental types of surveillance noise.

We prove for constrained mean reporting fractions and mean delays, that it is preferable to minimise variability among reporting fractions but to maximise the variability of the reporting delay distribution such that a minority of infections face large delays but the majority possess short lags to notification. This proceeds from standard *experimental design* theory applied to our metric, which shows that the information embedded within an epidemic curve depends on the product of the geometric means of the reporting fractions and cumulative delay probabilities corrupting that curve. This central result also provides a non-dimensional score for summarising and ranking the reliability of (or uncertainty within) different surveillance data for inferring pathogen transmissibility.

Last, we apply this framework to explore and critique a common claim in the literature, which asserts that death curves are more robust for inferring transmissibility than case curves. This claim, which as far as we can tell has never been formally verified, is usually made for acute infectious diseases such as COVID-19 and pandemic influenza [2, 20], where cases are severely under-reported, with symptom-based fluctuations in reporting. In such settings it seems plausible to reason that deaths are less likely under-counted and more reliable for *R*_*t*_ inference. However, we compute our metrics using COVID-19 reporting rate estimates [18, 26] and discover few instances in which death curves are definitively more informative or reliable than case counts.

While this may not rule out the possibility of having a more reliable death time series, it elucidates and exposes how the different noise terms within both data sources corrupt information and presents new methodology for exploring these types of questions more precisely. We illustrate how to compute our metrics practically using empirical COVID-19 and Ebola virus disease (EVD) noise distributions and outline how other common data such as hospitalisations, prevalence and wastewater virus surveys conform to our framework. Hopefully the tools we developed here will improve quantification of noise and information and highlight key areas where enhanced surveillance strategies can maximise impact.

## Results

### Methods overview

We summarise the salient points from the Methods and outline the main arguments that underpin all subsequent Results sections. Our analysis is centred on the *renewal model* [27], which is widely applied to describe the dynamics of epidemics of COVID-19, Ebola virus disease (EVD), influenza, dengue, measles, and many others [21]. This model posits that new infections at time step *t* (*I*_*t*_), are Poisson (Pois) distributed with mean that is the product of the instantaneous reproduction number (*R*_*t*_) and total infectiousness (Λ_*t*_). Here Λ_*t*_ defines how past infections engender new ones based on ***w***, the generation time distribution of the pathogen. In Eq. (15) and Table I we provide precise definitions of these variables.

**Table I:**
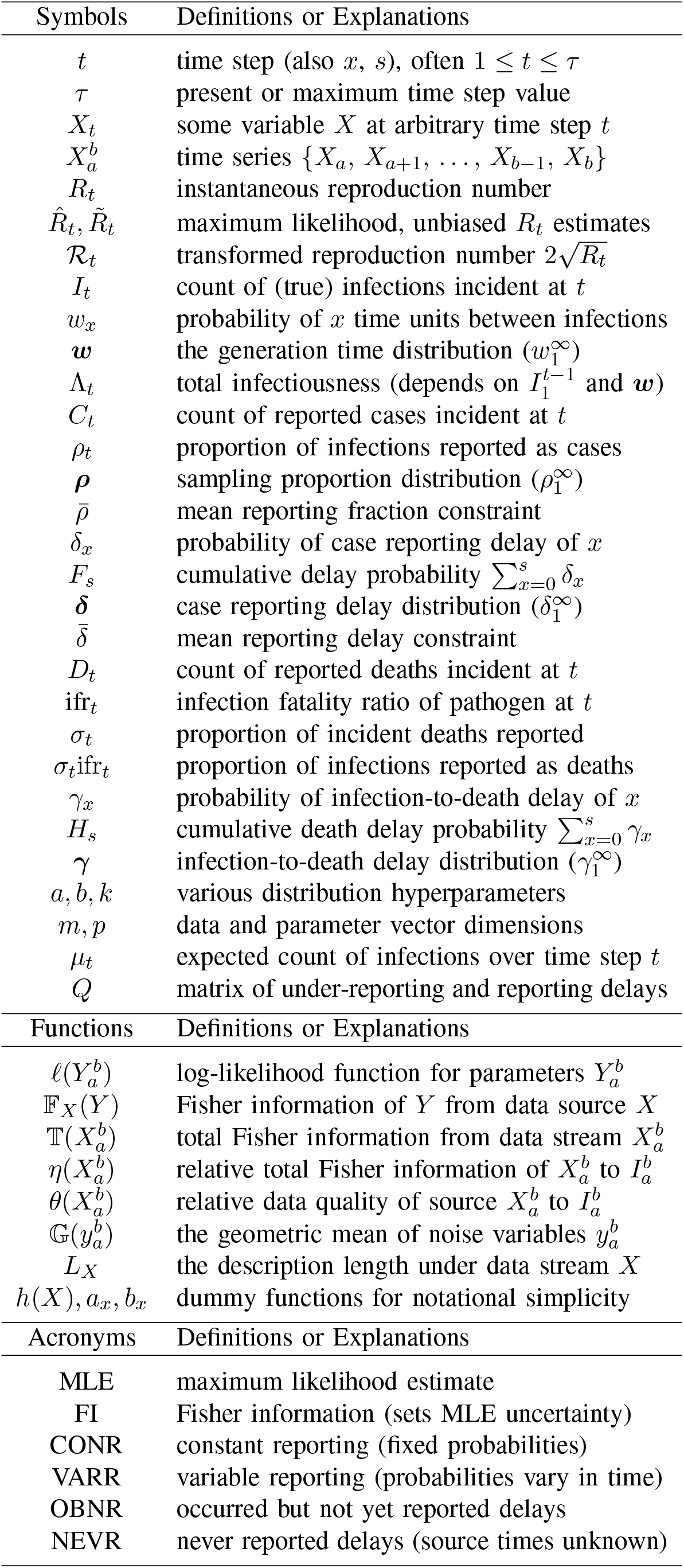
Summary of Notation.

An important problem in infectious disease epidemiology is the estimation of *R*_*t*_ across the duration of an epidemic [1]. However, as infections cannot be observed, we commonly have to infer *R*_*t*_ from noisy proxies such as the time series of reported cases or deaths. These can be described by generalised renewal models that include terms for practical noise sources such as under-reporting and delays in reporting [28]. We define these models in Eq. (1) and Eq. (2) and detail the properties of various noise sources in the Methods. Our aim is to understand and quantify how much information for inferring *R*_*t*_, as a fraction of what would be available if infections were observable, can be extracted from these proxies.

We pursue this aim by adapting concepts from statistical theory and information geometry. We first construct the log-likelihood function of the parameter vector 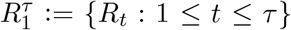, with *τ* as the present or last observation time and *t* scaled in units (e.g., weeks) so that each *R*_*t*_ can be assumed independent. This function is 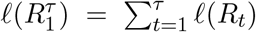 with *𝓁*(*R*_*t*_) := logℙ(*I*_*t*_ | *R*_*t*_) computed from the Poisson distribution of the renewal model. Eq. (16) results and admits the maximum likelihood estimates (MLEs), 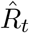 for all *t*, as its maxima. The reliability of these MLEs is characterised by the Fisher information (FI) of 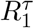 from the time series or curve of incident infections 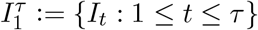.

Larger FI values imply smaller asymptotic uncertainty around the MLEs [29]. We obtain 𝔽_*I*_ (*R*_*t*_), the FI of *R*_*t*_, in Eq. (17) by evaluating the average curvature of the log-likelihood function. We then formulate the *total information*, 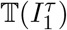, as a product of 𝔽_*I*_ (*R*_*t*_) terms across *t* as Eq. (18). This follows from the independence of the *R*_*t*_ variables and is a novel measure of the reliability of the infection time series. It is also delimits the maximum possible precision around the MLEs of *R*_*t*_ for any time series. Since 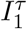 is often unobservable, 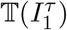 is generally not computable and a theoretical maximum. However, our subsequent results circumvent this issue.

In the upcoming sections we employ this same recipe of constructing a log-likelihood and computing MLEs and FI values but now for practical time series or data streams that are corrupted by under-reporting and delays. This yields Eq. (3)-Eq. (8), which contain the ingredients for deriving the total information in case, death and any other incidence data that is related to 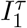 via a generalised renewal model (this includes prevalence, hospitalisations and virus abundance found in wastewater). We derive a key result for 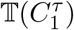 in Eq. (9), showing exactly how case data 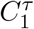 causes a loss in *R*_*t*_ estimate reliability.

Building on this expression we develop metrics 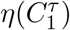 and 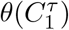 in Eq. (10)-Eq. (11), which effectively quan-tify 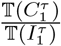 i.e., the level of informativeness of case data relative to true infections. The smaller these metrics are, the more information that is lost due to surveillance noise. Importantly, these metrics are analytic, require no knowledge of 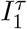 or the generation time distribution (both are difficult to observe) and are interpretable since each noise type contributes a separate geometric mean term. Further, they play an integral role in defining the statistical complexity of the generalised renewal model describing that time series, as we find in Eq. (12).

Repeating the above recipe we obtain similar metrics for death data 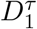, by characterising the ratio 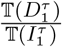 in Eq. (13)-Eq. (14). We can similarly compute ratios for hospitalisations, prevalence and viral wastewater data by including appropriate delay and under-reporting terms as needed. We complete our results by including empirical estimates of case and death noise sources within our framework to compare 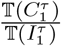 and 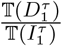 for COVID-19 and EVD. We find that the common assumption of deaths being more informative than cases is not likely true for COVID-19 but holds under some conditions for EVD.

### Renewal models with noisy observations

We denote the empirically observed or reported number of cases at time step or unit *t*, subject to noise from both under-reporting and reporting delays, as *C*_*t*_ with 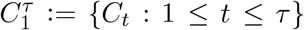 as the epidemic case curve. This curve is obtained from routine outbreak surveillance and is a corrupted version of the true incidence 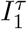 [10], modelled by Eq. (15). These noise sources (see Methods for statistical descriptions) are parametrised by reporting fractions 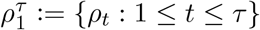 and a delay distribution ***δ*** := {*δ*_*x*_ : *x* ≥ 0}. Here *ρ*_*t*_ is the fraction of infections reported as cases at *t* and the *δ*_*x*_ the probability of a lag from infection times to case report of *x* units.

We assume that these noise sources are estimated from auxiliary line-list or contact tracing data [12, 30]. As a result, we can construct Eq. (1) as in [25] (see Methods). Note that if noise source estimates are unavailable then 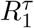 becomes statistically non-identifiable or ill-defined.

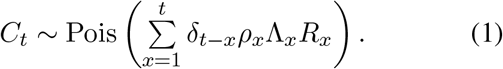

This noisy renewal model suggests that *C*_*t*_ (unlike *I*_*t*_) contains partial information about the entire time series of reproduction numbers for *x* ≤ *t* as mediated by delay and reporting probabilities. Perfect reporting corresponds to *ρ*_*x*_ = 1 for all *x, δ*_0_ = 1 (*δ*_*x* ≠0_ = 0) and means *C*_*t*_ →*I*_*t*_. The models in (i)-(ii) of the Methods are obtained by individually removing noise sources from Eq. (1).

Other practical epidemic surveillance data such as the time series of new deaths or hospitalisations conform to the framework in Eq. (1) either directly or with additional effective delay and under-reporting stages [20]. The main one we investigate here is the count of new deaths (due to infections) across time, which we denote 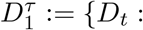 1 ≤ *t* ≤ *τ*}. The death curve involves a reporting delay that includes the intrinsic lag from infection to death. We let ***γ*** := {*γ*_*x*_ : *x* ≥ 0} represent the distribution of lag from infection to observed death and 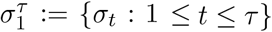 be the fraction of deaths that are reported.

An important additional component when describing the chain from 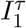 to 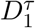 is the infection fatality ratio,ifr_*t*_, which is the probability at time *t* that an infection culminates in a death event [10]. Fusing these components yields Eq. (2) as a model for death counts *D*_*t*_.

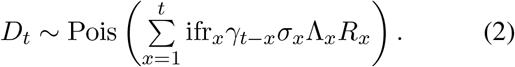

In a later section we explain how analogues of Eq. (1)-Eq. (2) also fit other data streams such as hospitalisations and prevalence. Some studies [2, 31] replace this Pois formulation with a negative binomially (NB) distribution to model extra variance in these data. In the Appendix we show that this does not disrupt our subsequent results on the relative informativeness of surveillance data (though the NB formulation is less tractable and unsuitable for extracting generalisable, simulation-free insights).

### Fisher information derivations for practical data

We derive the FI of parameters 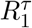 given the case curve 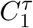. This procedure mirrors that used in the Methods to obtain Eq. (18). We initially assume that reporting delays are OBNR i.e., that we eventually learn the source time of cases at a later date. This corresponds to a right censoring that can be compensated for using *nowcasting* techniques [13]. Later we prove that this not only defines a practical noise model but also serves as an upper bound on the information available from NEVR delays, where the true timestamps of cases are never resolved. Mathematically, the OBNR assumption lets us decompose the sum in Eq. (1). We can therefore identify the component of *C*_*t*_ that is informative about *R*_*x*_. This follows from the statistical relationship *C*_*t*_ | *R*_*x*_ ∼ Pois (*δ*_*t*−*x*_*ρ*_*x*_Λ_*x*_*R*_*x*_).

As we are interested in the total information that 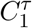 contains about every *R*_*t*_ we collect and sum contributions from every *C*_*t*_. We can better understand this process by constructing the matrix *Q* in Eq. (3), which expands the convolution of the reporting fractions with the delay probabilities over the entire observed time series.

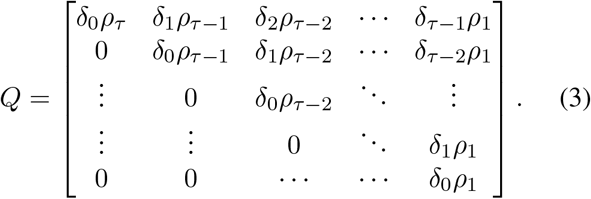

We work with the vector *µ* = [*µ*_*τ*_, *µ*_*τ*−1_, …, *µ*_1_]^⊤^ with *µ*_*t*_ = Λ_*t*_*R*_*t*_ and ⊤ denoting the transpose operation. Then *Qµ* = [𝔼[*C*_*τ*_], 𝔼[*C*_*τ*−1_], …, 𝔼[*C*_1_]]^⊤^ with 𝔼[*C*_*t*_] as the mean of the reported case incidence at time *t*.

The components of 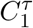 that contain information about every *R*_*t*_ parameter follow from *Q*^⊤^*µ* = [*δ*_0_*ρ*_*τ*_ *µ*_*τ*_, (*δ*_0_ + *δ*_1_)*ρ*_*τ*−1_*µ*_*τ*−1_, …, (*δ*_0_ + … + *δ*_*τ*−1_)*ρ*_1_*µ*_1_]. The elements of this vector are Poisson means formed by collecting and summing the components of 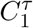 that inform about [*R*_*τ*_, *R*_*τ*−1_, …, *R*_1_], respectively. Hence we obtain the key relationship in Eq. (4) with 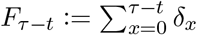 as the cumulative probability delay distribution.

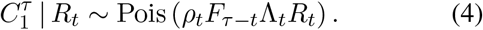

The ability to decompose the row or column sums from *Q* into the Poisson relationships of Eq. (4) is a consequence of the independence properties of renewal models and the infinite divisibility of Poisson formulations.

Using Eq. (4) and analogues to Poisson log-likelihood definitions from the Methods we derive the FI that 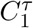 contains about *R*_*t*_ as follows in Eq. (5).

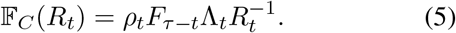

As in Eq. (17) we recompute the FI in Eq. (5) under the transform 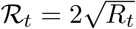 to obtain 𝔽_*C*_(ℛ _*t*_) = *ρ*_*t*_*F*_*τ*−*t*_Λ_*t*_. It is clear that under-reporting and delays can substantially reduce our information about instantaneous reproduction numbers. As we might expect, if *ρ*_*t*_ = 0 (no reports at time unit *t*) or *F*_*τ*−*t*_ = 0 (all delays are larger than *τ* − *t*) then we have no information on *R*_*t*_ at all from 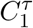. If reporting is perfect then *ρ*_*t*_ = 1, *F*_*τ*−*t*_ = 1 and 𝔽_*C*_(*R*_*t*_) is equal to the FI from 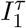 in Eq. (17).

The MLE, 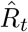, also follows from Eq. (4) (see Methods) as 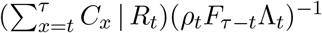, with *C*_*x*_ | *R*_*t*_ as the component of *C*_*x*_ containing information about *R*_*t*_. By comparison with the MLE under perfect surveillance we see that 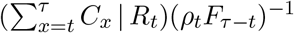 is equivalent to applying a nowcasting correction as in [12, 13]. An important point to make here is that while such corrections can remove bias, allowing inference despite these noise sources, they cannot improve on the information (in this case Eq. (5)) inherently available from the data. This is known as the data processing inequality [32, 33].

If we cannot resolve the components of every *C*_*t*_ from Eq. (1) as 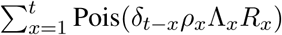, then the re-porting delay is classed as NEVR (i.e., we never uncover case source dates). Hence we know *Qµ* but not *Q*^⊤^*µ*. Accordingly, we must use Eq. (1) to construct an aggregated log-likelihood 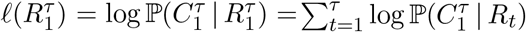 gives Eq. (6) with the aggregate term 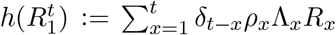. We ignore constants that do not depend on any *R*_*t*_ in this likelihood.

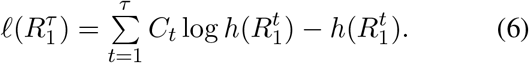

For every given *R*_*t*_ we decompose 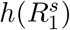 for *s* ≥ *t* into the form *δ*_*s*−*t*_*ρ*_*t*_Λ_*t*_*R*_*t*_+*a*_*t*_, where *a*_*t*_ collects all terms that are not informative about that specific *R*_*t*_. Here *s ≥ t* simply indicates that information about *R*_*t*_ is distributed across later times due to the reporting delays.

We can then obtain the FI contained in 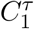 about *R*_*t*_ by computing 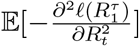, yielding Eq. (7) (see Appendix for derivation details), with *b*_*t*_ := *a*_*t*_(*δ*_*s*−*t*_*ρ*_*t*_Λ_*t*_*R*_*t*_)^−1^.

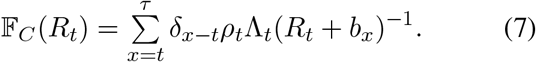

If we could decouple the interactions among the reproduction numbers then the *b*_*x*_ terms would disappear and we would recover the expressions derived under OBNR delay types. Since *b*_*x*_ is a function of other reproduction numbers, the overall FI matrix for 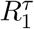 is not diagonal (there are non-zero terms from evaluating 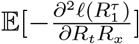).

However, we find that this matrix can be reduced to a triangular form with determinant equal to the product of terms (across *t*) in Eq. (7). We show this for the example scenario of *τ* = 3 in the Appendix. As a result, the FI term for *R*_*t*_ in Eq. (7) does behave like and correspond to that in Eq. (5). Interestingly, as *b*_*x*_ ≥ 0, Eq. (7) yields the revealing inequality 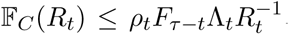. This proves that OBNR delays upper bound the information available from NEVR delays. Last, we note that robust transforms cannot be applied to remove the dependence of Eq. (7) on the unknown *R*_*t*_ parameters. The best we can do is evaluate Eq. (7) at the MLEs 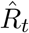, for all *t*.

These MLEs emerge as the joint maxima of the set of coupled differential equations 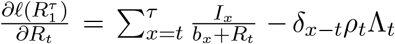 i.e., numerical solutions of Eq. (8) for all *t*.

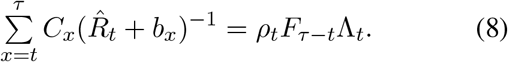

Here sums start at *t* as they include only time points that contain information about *R*_*t*_. Expectation-maximisation algorithms, such as the *deconvolution* approaches outlined in [10], are viable means of computing these MLEs or equivalents. Note that the nowcasting methods used to correct for OBNR delays do not help here [12] and that for both OBNR and NEVR delays the cumulative probability terms must be aggregated to match chosen time units (e.g., if empirical delay distributions are given in days but *t* is in weeks then *F*_*x*_ sums over 7*x* days).

### Reliability measures for surveillance data

Having derived the FI for each instantaneous reproduction number, we provide a measure of the total information that 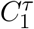 provides about 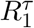 or the transformed 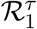. As detailed in the Methods, this total information, 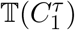, relates inversely to the smallest joint uncertainty around unbiased estimates of all our parameters [34]. As larger 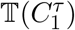 implies reduced overall uncertainty, this is a rigorous measure of the statistical reliability of noisy data sources for inferring pathogen transmissibility. Use of this or related metrics for quantifying the information in noisy epidemic data is novel (as far as we can tell).

We first consider the OBNR delay case under arbitrarily varying (VARR) reporting rates. Since the FI matrix under OBNR delays is diagonal, with each element given by Eq. (5), we can adapt Eq. (18) to derive Eq. (9).

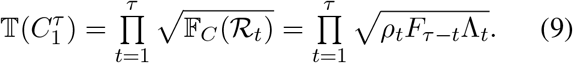

Here we have applied the 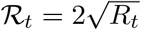 transformation to show that the total information in this noisy stream can be obtained without knowing *R*_*t*_. In the absence of this transform we would have 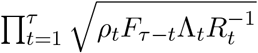.

Since 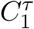 is a distortion of the true infection incidence 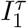 we normalise Eq. (9) by Eq. (18) to develop a new reliability metric, 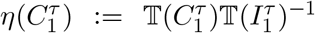. This is given in Eq. (10) and valid under both *R*_*t*_ and ℛ _*t*_.

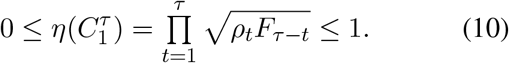

We can relate this reliability measure to a fixed, effective reporting fraction, 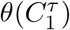, which causes an equivalent information loss. Applying Eq. (10), we get 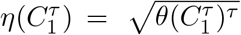, which yields Eq. (11). Here 𝔾(.) indicates the *geometric mean* of its arguments over 1 ≤ *t* ≤ *τ*.

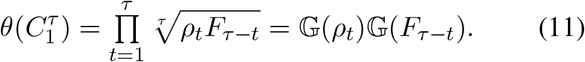

Eq. (11) is a central result of this work. It states that the total information content of a noisy epidemic curve is independently modulated by the geometric mean of its reporting fractions, 𝔾(*ρ*_*t*_), and that of its cumulative delay probabilities, 𝔾(*F*_*τ*−*t*_). Moreover, Eq. (11) provides a framework for gaining analytic insights into the separate influences of both noise sources from different surveillance data and for ranking the overall quality of those diverse data. For example, we immediately see that 𝔾(.) is bounded by the smallest and largest noise term across *t*. Importantly, Eq. (11) has no dependence on Λ_*t*_, which is generally unknown and sensitive to difficult-to-infer changes in the generation time distribution [35].

Eq. (11) applies to OBNR delays exactly and upper bounds the reliability of data streams with NEVR delays (see previous section). Tractable results for NEVR delays are not possible and necessitate numerical computation of Hessian matrices of 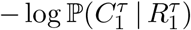 (we outline the log-likelihoods and other equations for a *τ* = 3 example in the Appendix). However, we find that the Eq. (11) upper bound is tight for two elementary settings. The first is under a constant or deterministic delay of *d* i.e., *δ*_*x*=*d*_ = 1. Eq. (1) reduces to *C*_*t*_ ∼ Pois(*ρ*_*t*−*d*_Λ_*t*−*d*_*R*_*t*−*d*_). As each *C*_*t*_ only informs on *R*_*t*−*d*_ OBNR and NEVR delays are the same and corrected by truncation. Degenerate delays such as these can serve as useful elements for constructing complex distributions [36].

The second occurs when transmissibility is constant or stable i.e., *R*_*t*_ = *R* for all *t*. This applies to inferring the basic reproduction number (*R*_0_) during initial phases of an outbreak [1]. We can sum Eq. (5) to get 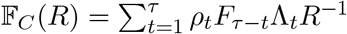 for OBNR delays. We can calculate the FI for NEVR delays from Eq. (6), which admits a derivative 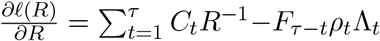 and hence a FI and MLE that are precisely equal to those for OBNR delays. This proves a convergence in the impact of two fundamentally different delay noise sources and emphasises that noise has to be contextualised with the complexity of the signal to be inferred. Simpler signals, such as a stationary *R* that remains robust to the shifts and reordering of 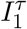 due to delays, may be notably less susceptible to fluctuations in noise probabilities.

### Ranking noise sources by their information loss

The metric proposed in Eq. (11) provides an original and general framework for scoring proxies of incidence (e.g., epidemic case curves, death counts, hospitalisations and others) using only their noise probabilities and without the need for simulations. We explore the implications of Eq. (11) both for understanding noise and ranking those proxies. The geometric mean decomposition allows us to separately dissect the influences of under-reporting and delays. We start by applying experimental design theory [37, 38], to characterise the best and worst noise types for inferring effective reproduction numbers.

We consider 𝔾(*ρ*_*t*_), the geometric mean of the reporting probabilities across time. If we assume the average sampling fraction 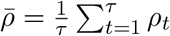 is fixed (e.g., by some overall surveillance capacity) then we immediately know from design theory that 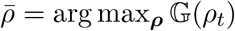. This means that of all the possible distributions of sampling fractions fitting that constraint, ***ρ***, CONR or constant reporting with probability 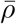 is the most informative [39]. This result is new but supports earlier studies recognising that CONR is preferred to VARR, although they investigate estimator bias and not information loss [9, 21].

Accordingly, we also discover that the worst sampling distribution is maximally variable. This involves setting *ρ*_*t*_ ≈ 1 for some time subset 𝕊 such that 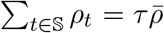 with all other *ρ*_*t*_ ≈ 0 (we use approximate signs as we assume non-zero sampling probabilities). Relaxing this constraint, Eq. (11) presents a framework for comparing different reporting protocols. We demonstrate these ideas in Fig. 2, where *ρ*_*t*_ ∼ Beta(*a, b*) i.e., each reporting fraction is a sample from a Beta distribution. Reporting protocols differ in (*a, b*) choices. We select 10^4^ *ρ*_*t*_ samples each from 2000 distributions with 10^−1^ ≤*b* ≤ 10^2^ and *a* computed to fulfil the mean constraint 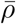. Variations in the resulting 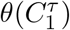 metrics indicate the influence of reporting fraction uncertainties under this mean.

**Fig. 2:**
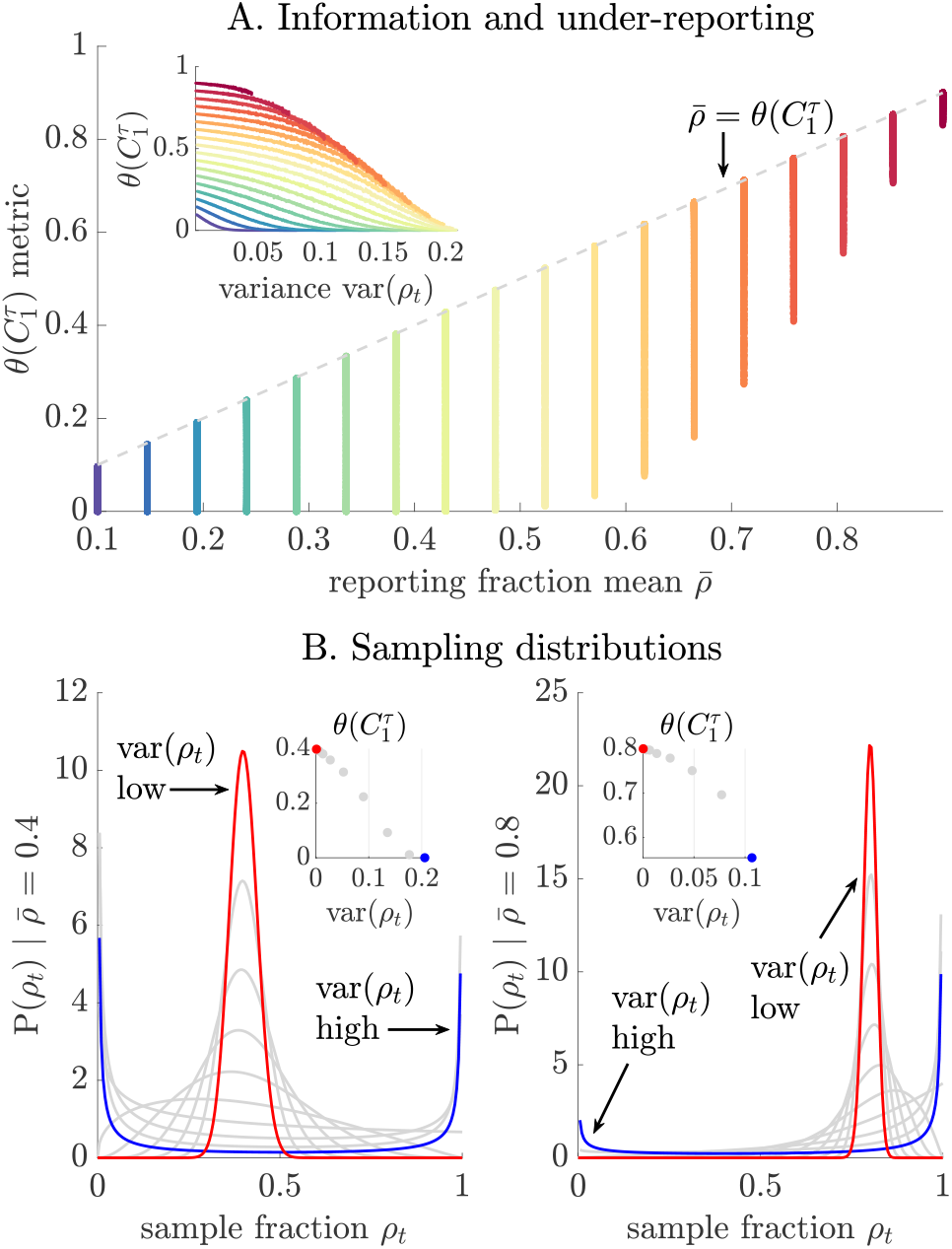
The information loss in under-reporting. We investigate the effective information metric 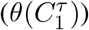 for variable reporting strategies (VARR) with reporting fraction *ρ*_*t*_ drawn from various Beta distributions. Panel A shows that while 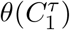 depends on the mean reporting probability 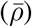, large fluctuations in the total information can emerge from the level of variability, controlled here by the Beta distribution shape. This follows from the overlap of the coloured curves, which compute 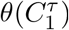 at fixed 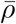. The grey line (dashed) is the optimal constant reporting (CONR) protocol. Our metric decreases with the variance of the protocol (var(*ρ*_*t*_)) as seen in the inset with matching colours for each 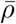. Panel B illustrates the Beta sampling distributions and their resulting variance and metric scores (inset). The most variable reporting strategy (blue) is the worst protocol for a given 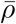.

Panel A of Fig. 2 shows that 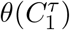 generally increases with the mean reporting probability 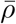. However, this improvement can be denatured by the variance, var(*ρ*_*t*_), of the reporting scheme (inset where each colour indicates the various schemes with a given 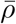). The CONR scheme is outlined with a grey line (dashed) and, as derived, is the most informative. Panel B confirms our theoretical intuition on how var(*ρ*_*t*_) reduces total information with the extreme (worst) sampling scheme outlined above in blue and the most stable protocol in red. There are many ways to construct *ρ*_*t*_ protocols. We chose Beta distributions because they can express diverse reporting probability shapes using only two parameters.

Similarly we investigate reporting delays via 𝔾(*F*_*τ*−*t*_), the geometric mean of the cumulative delay or latency distribution across time. Applying a mean delay constraint 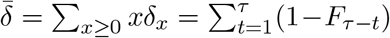 (e.g., reflecting operational limits on the speed of case notification), we adapt experimental design principles. As we effectively maximise a FI determinant (see derivation of Eq. (11)) our results are termed *D-optimal* [39]. These suggest that max_***δ***_ 𝔾(*F*_*τ*−*t*_) is achieved by cumulative distributions with the most uniform shape. These possess the largest *δ*_0_ within this constraint. Delay distributions with significant dispersion (e.g., heavy tails) attain this optima while fixed delays (where 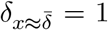 and 0 otherwise) lead to the largest information loss under this constraint.

This may seem counter-intuitive as deterministic delays best preserve information outside of that delay and can be treated by truncating the observed epidemic time series e.g., for a fixed weekly lag we can ignore the last week of data. However, this causes a bottleneck. No information is available for that truncated week eliminating any possibility of timely inference (and making epidemic control difficult [40]). In contrast, a maximally dispersed delay distribution slightly lags the majority of cases, achieving the mean constraint with large latencies on a few cases. This ensures that, overall, we gain more actionable information about the time series.

We illustrate this point (and relax the mean constraint) in Fig. 3, where we verify the usefulness of Eq. (11) as a framework for comparing the information loss induced by delay distributions of various shapes and forms. We model ***δ*** as 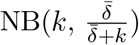 with *k* describing the dispersion of the delay. Panel A demonstrates how our 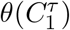 metric varies with *k* (30 values taken between 10^−1^ and 10^2^) at various fixed mean constraints (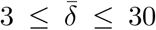, each given as a separate colour). In line with the theory, we find that decreasing *k* (increasing dispersion of the delay distribution) improves information at any given 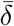.

**Fig. 3:**
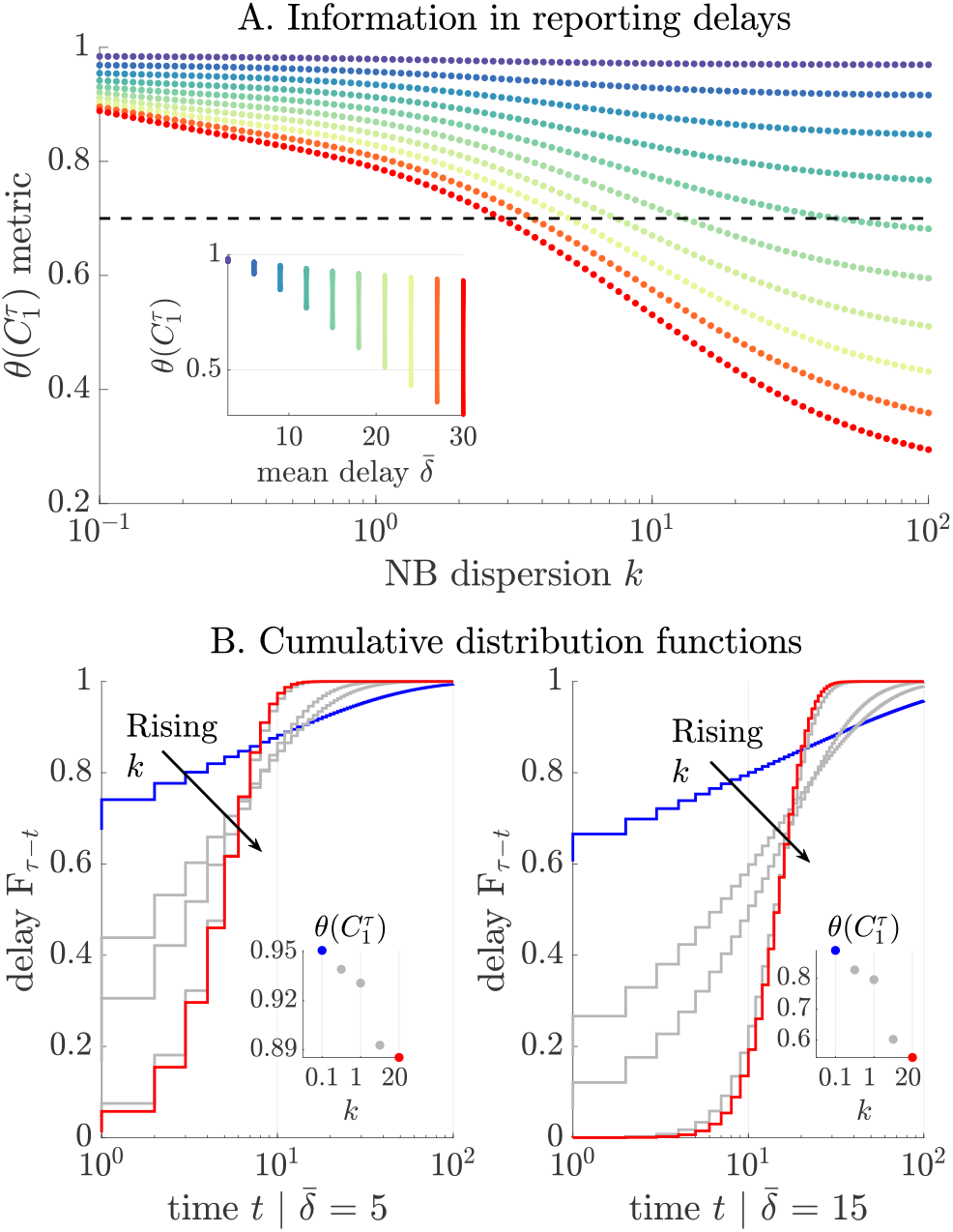
The information loss from delays. We compute the information metric 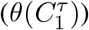 for various delay distributions, which are negative binomial (NB) with some dispersion *k*. Panel A examines how mean delay 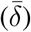 and *k* influence the information loss, showing that various combinations can result in the same loss (intersections of the coloured curves, each representing a different 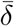, with the dashed black line). Further, the inset illustrates the variations in our metric at a given mean (matching colours) due to the shape of the delay distribution. Panel B confirms this relationship and indicates that the most dispersed distributions (smallest *k*, blue, with largest start to cumulative delay distribution *F*_*τ*−*t*_) preserve the most information as compared to more deterministic delays (red, largest *k*). The insets verify this point.

The importance of both the shape and mean of reporting delays is indicated in the inset as well as by the number of distributions (seen as intersects of the dashed black line) that result in the same 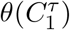. Panel B plots corresponding cumulative delay probability distributions, validating our assertion from design theory that the best delays (blue, with metric in inset) are dispersed, forcing *F*_*τ*−*t*_ high very early on (maximise *δ*_0_ and leading to the most uniform shape), while the worst ones are more deterministic (red, larger *k*). These curves are for OBNR delays and upper bound the performance expected from NEVR delays except for the settings described in the previous section where both types coincide.

### Comparing different epidemic data streams

Our metric (Eq. (11)) not only allows the comparison of different under-reporting schemes and reporting delay protocols (see above section) but also provides a common score for assessing the reliability or informativeness of diverse data streams for inferring 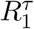. The best stream, from this information theoretic viewpoint, maximises the product of the geometric means 𝔾(.) of the cumulative delay probabilities *F*_*τ*−*t*_ and reporting fractions *ρ*_*t*_. Many common surveillance data types used for inferring pathogen transmissibility have been modelled within the framework of Eq. (1) and therefore admit related *θ*(.) metrics. Examples include time series of deaths, hospitalisations, the prevalence of infections and incidence proxies generated from viral surveys of wastewater.

We detail death count data in the next section but note that its model, given in Eq. (2), is a simple extension of Eq. (1). Hospitalisations may be described similarly with the ifr term replaced by the proportion of infections hospitalised and the intrinsic delay distribution defining the lag from infection to hospital admission [1]. The infection prevalence conforms to Eq. (1) because it can be represented as a convolution of the infections with a duration of infectiousness distribution, which essentially contributes a reporting delay [41]. Viral surveys also fit Eq. (1). They offer a downsampled proxy of incidence, which is delayed by a shedding load distribution defining the lag before infections are detected in wastewater [42]. Consequently, our metrics are widely applicable.

While in this study we focus on developing methodology for estimating and contrasting the information from the above surveillance data we find that our metric is also important for defining the complexity of a noisy renewal epidemic model. Specifically, we re-derive Eq. (11) as a key term of its *description length* (*L*). Description length theory evaluates the complexity of a model from how succinctly it describes its data (e.g., in bits) [34, 43]. This measure accounts for model structure and data quality and admits the approximation 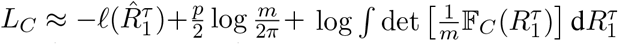. Here the first term indicates model fit by assessing the log-likelihood at our MLEs 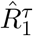. The second term includes data quality through the number of parameters (*p*) and data size (*m*). The final term defines how model structure shapes complexity with the integral across the parameter space of 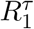.

This formulation was adapted for renewal model selection problems in [44] assuming perfect reporting. We extend this and show that our proposed total information 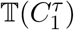 plays a central role. Given some epidemic curve 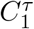 we can rewrite the previous integral as 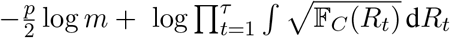 and observe that *m* = *p* = *τ*. It is known that under a robust transform such as 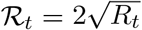 this integral is c onserved [34, 38]. Consequently, 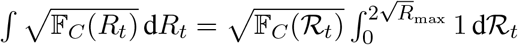 with *R*_max_ as some maximum value that every *R*_*t*_ can take. Combining these expressions we obtain Eq. (12), highlighting the importance of our total information metric.

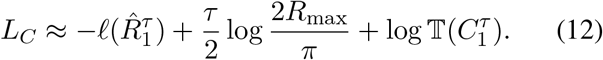

If we have two potential data sources for inferring 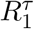 then we should select the one with the smaller *L*_*C*_ value. Since the middle term in Eq. (12) remains unchanged in this comparison, the key points when comparing model complexity relate to the level of fit to the data and the total Fisher information of the model given that data [43]. Using Δ to indicate differences this comparison may be formulated as 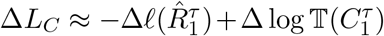. The second term can be rewritten as 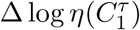 (see Eq. (10)). This signifies that these metrics play a central role when comparing different data streams.

### Are COVID-19 deaths or cases more informative?

In the above sections we developed a framework for comparing the information within diverse but noisy data streams. We now apply these results to better understand the relative reliabilities of two popular sources of information about transmissibility 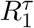 ; the time series of new cases 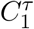 and of new death counts 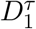. Both data streams have been extensively used across the ongoing COVID-19 pandemic to better characterise pathogen spread [1]. Known issues stemming from fluctuations in the ascertainment of COVID-19 cases [18, 19] have motivated some studies to assert 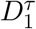 as the more informative and hence trustworthy data for estimating 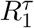 [2, 20].

These works have reasonably assumed that deaths are more likely to be reliably ascertained. Case reporting can be substantially biased by testing policy inconsistencies and behavioural changes (e.g., symptom based healthcare seeking). In contrast, given their severity, deaths should + be less likely to be under-ascertained [1]. However, no analysis, as far as we are aware, has explicitly tested this assumption. Here we make some progress towards better comprehending the relative merits of both data streams. We start by computing ratios of our metric in Eq. (11) for both 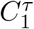 and 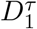 via Eq. (1) and Eq. (2).

This results in 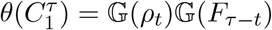 for cases and, by analogy, 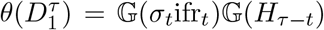 for deaths. In the same way that *ρ*_*t*_ defines the proportion of infections reported as cases, the product *σ*_*t*_ifr_*t*_ defines the proportion of infections that are reported as deaths. This follows as ifr_*t*_ is the fraction of infections that engender deaths and *σ*_*t*_ is the proportion of those deaths that are reported. While *F*_*τ*−*t*_ is the cumulative probability of reporting delays up to *τ* −*t* time units, 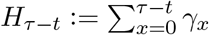 describes the cumulative probability of delays from infection to death up to *τ* − *t* time units in duration.

Using shorthand 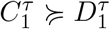 for when 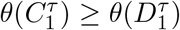 i.e., ≽ indicates greater than or equals with respect to total information, we obtain Eq. (13). We rearrange terms to get reporting fractions and delays on different sides by decomposing the geometric mean of a product into products of the geometric means in each term.

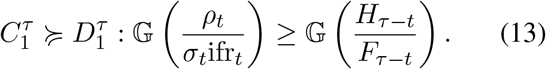

Eq. (13) states that cases are more informative when the geometric mean of the case to death reporting fractions is at least as large as that of the death and case cumulative delays. Studies preferring death data effectively claim that the variation in case reporting probabilities *ρ*_*t*_ (which we proved in a previous section always decreases the geometric mean for a given mean constraint) is sufficiently strong to mask the influences of the infection fatality ratio (ifr_*t*_), the death reporting probability (*σ*_*t*_) and any expected variations in those quantities.

Proponents of using death data to infer 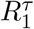 recognise that the infection to death delay (with cumulative distribution *H*_*τ*−*t*_) is appreciably larger in mean than that of corresponding reporting lags from infection (*F*_*τ*−*t*_) and therefore unsuitable for real time estimation (where this extra lag denatures recent information as we showed in earlier sections). We allow for all of these adjustments. We assume that the infection fatality ratio is constant at ifr (maximising 𝔾(ifr_*t*_)) and that death ascertainment is perfect (*σ*_*t*_ = 1). Even for purely retrospective estimation with correction for delays we expect 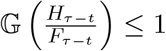. We set this to 1, maximising the informativeness of 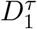.

Combining these assumptions we reduce Eq. (13) into Eq. (14). This presents a sufficient condition for case data to be more reliable than the death time series.

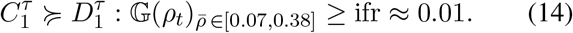

Here we choose a relatively large ifr for COVID-19 of 1% [45]. Case reporting fraction estimates range from about 7% to 38% [18], which we apply to constrain 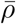, the average *ρ*_*t*_. Inputting these estimates, we examine possible *ρ*_*t*_ sampling distributions under the Beta(*a, b*) formulation from earlier sections. Our main results are in Fig. 4. We take 10^4^ samples of *ρ*_*t*_ from each of 2000 distributions parametrised over 10^−1^ ≤*b* ≤ 10^2^ with *a* set to satisfy our mean 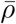 reporting constraints.

**Fig. 4:**
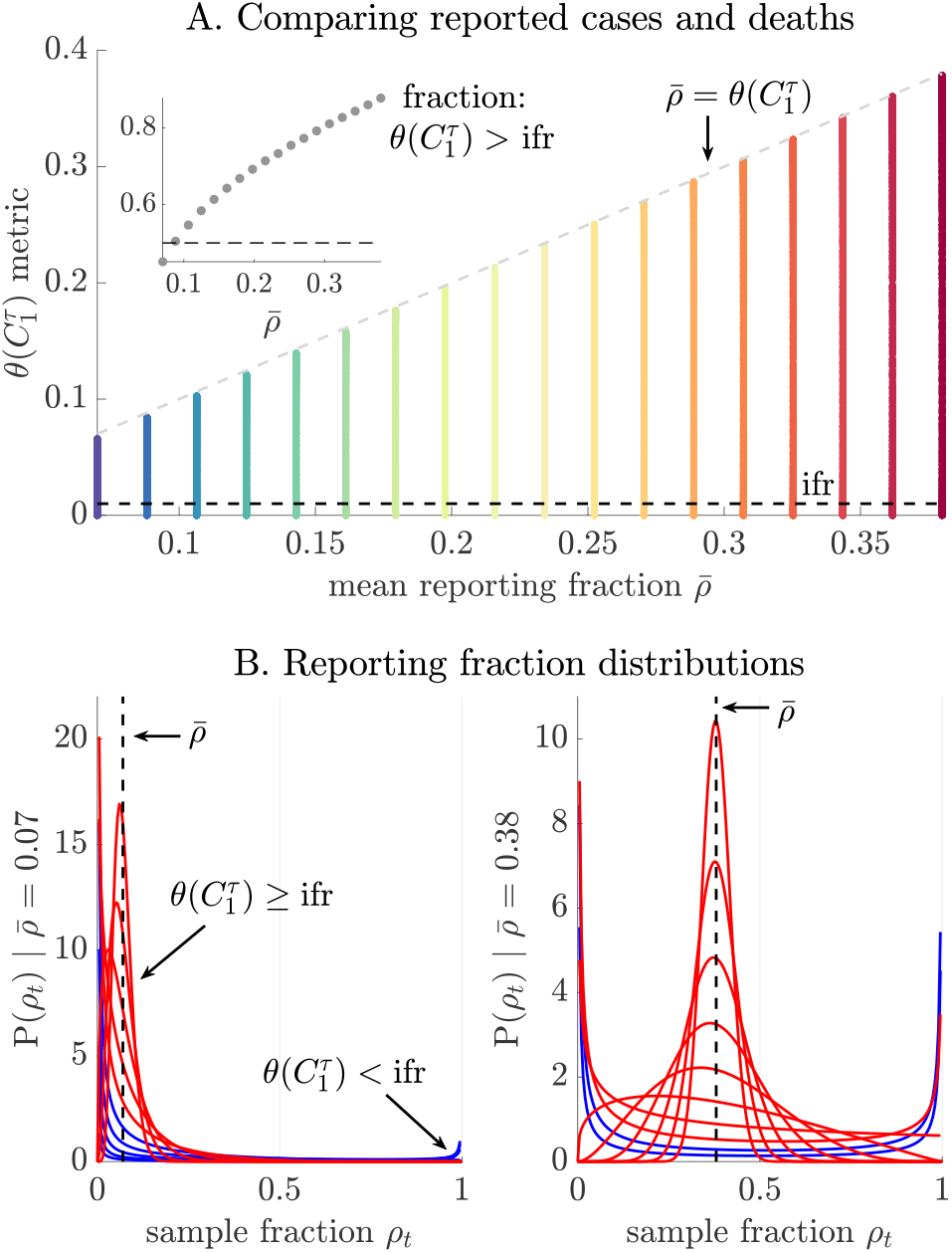
Epidemic case data may be more informative than death counts. Using metrics 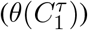 we compare the information in case curves 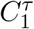 and death counts 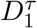 under assumptions that lead to Eq. (14). We examine various reporting strategies parametrised as Beta distributions with means 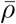 from 0.07 to 0.38 [18] and compare the resulting 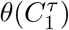 against the equivalent from deaths (which reduces to just the infection fatality ratio, ifr). Panel A shows that many such distributions for sampled cases (each colour considers a fixed 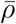) still contain more information than available from deaths (proportion of vertical lines above the black dashed threshold, plotted inset). Panel B plots those distributions at the ends of the empirical 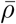 range with red indicating when 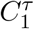 is more reliable. Substantial fluctuations in 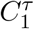 reporting can still preserve more information than might be found in 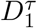.

Panel A plots our metric against those constraints (a different colour for each 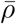) and the ifr threshold (black dashed). Whenever 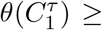 ifr we find that case data are more reliable. This appears to occur for many possible combinations of *ρ*_*t*_. The inset charts the proportion of Beta distributions that cross that threshold. This varies from about 45% at 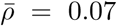 to 90% at 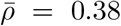. While these figures will differ depending on how likely a given level of variability is, they offer robust evidence that death counts are not necessarily more reliable. Even when deaths are perfectly ascertained (*σ*_*t*_ = 1) the small ifr term in 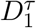 means that 99% of the original incidence data is lost, contributing appreciable uncertainty.

These points are reinforced by the design choices we have made, which inflate the relative information in the death time series. In reality *σ*_*t*_ *<* 1, ifr *<* 0.01, neither is constant [45, 46] and the uncertainty we include around 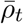 is wider than that inferred in [18]. Our results are therefore resilient to uncertainties in noise source estimates. Panel B displays the distributions of our sampling fractions with red (blue) indicating which shapes provide more (less) information than death data (see Eq. (14)). Our results also hold for both real time and retrospective analyses as we ignored the noise induced by the additional delays that death data contain (relative to case reports) when we maximised 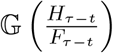.

Consequently, death data cannot be assumed, without rigorous and context-specific examination, to be generally more epidemiologically meaningful. For example, while 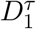 is unlikely to be more reliable in well-mixed populations, it may be in high-risk settings (e.g., care homes) where the local ifr is notably larger. Vaccines and improved healthcare, which substantially reduce ifr values in most contexts, will make death time series less informative about 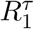. However, pathogens such as Ebola virus, which induce large ifr parameters, might result in death data that are more reliable than their case counts. We explore these points and demonstrate the practical applicability of our metrics in the next section.

### Practical applications of information metrics

Our metrics provide an interpretable, simulation agnostic and easily computable approach to quantifying the relative reliability of different epidemic time series. Because *θ*(.) is independent of usually unknown *R*_*t*_ and Λ_*t*_ terms it is robust to generation time misspecification and only requires estimates of noise terms for its calculation (hence no epidemic curve simulations are needed). Moreover, it depends purely on the geometric means of noise variables, which can be decomposed such that the influence of any noise source is clearly interpreted from the magnitude of its specific mean (see Eq. (11)).

These properties make *θ*(.) practically useful and we illustrate the benefits of our methodology using COVID-19 and EVD examples. In contrast to Fig. 4 where we maximised the information in deaths and minimised that from cases to bolster our rejection of the assertion that death data are definitively more informative, here we focus on inputting empirical noise distributions derived from real data. When distributions are unavailable we describe noise uncertainties via maximum entropy distributions based on what estimates are available (e.g., these are geometric, Geo, if a mean is given and uniform, Unif, over 95% credible intervals).

For COVID-19 we once again examine if death data are more reliable. From Eq. (13) we conclude 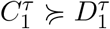 if 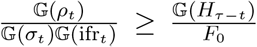. This follows as *F*_0_ = *δ*_0_ = min 𝔾(*F*_*τ*−*t*_) and ensures (if we are using NEVR delays) that we do not take ratios of upper bounds as 𝔾(*H*_*τ*−*t*_) already bounds the information in the infection-to-death delay. If delays are OBNR then Eq. (13) will be exact. We model *ρ*_*t*_ ∼ Unif(0.06, 0.08) [18], 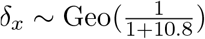 [47], 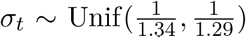 [46], 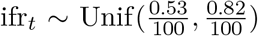 [45], 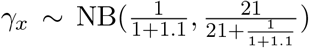 [48] and sample from these distributions 10^4^ times. We compute the terms in the inequality above and represent the relative information as 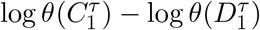 for easy visualisation.

This leads to the top panel of Fig. 5. Despite our use of the smallest reporting proportions from [18] we find that death data are less reliable. For EVD, we test the alternative hypothesis that case data are less reliable in the bottom panel of Fig. 5. We decide 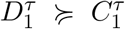 if 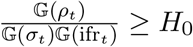 as we know *H*_0_ = *γ*_0_ = min 𝔾(*H*_*τ*−*t*_) and max 𝔾(*F*_*τ*−*t*_) = 1. We let *σ*_*t*_ = 1 (no estimates were easily available) and model *ρ*_*t*_ ∼ Unif(0.33, 0.83) [16], ifr_*t*_ Unif(0.69, 0.73) [49] and 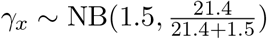 (roughly from [49]). The negative values of 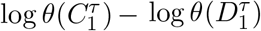 in Fig. 5 suggest EVD death data as the more informative source. However, this can change if *σ*_*t*_ *≪*1 as the difference is not as strong as for COVID-19.

**Fig. 5:**
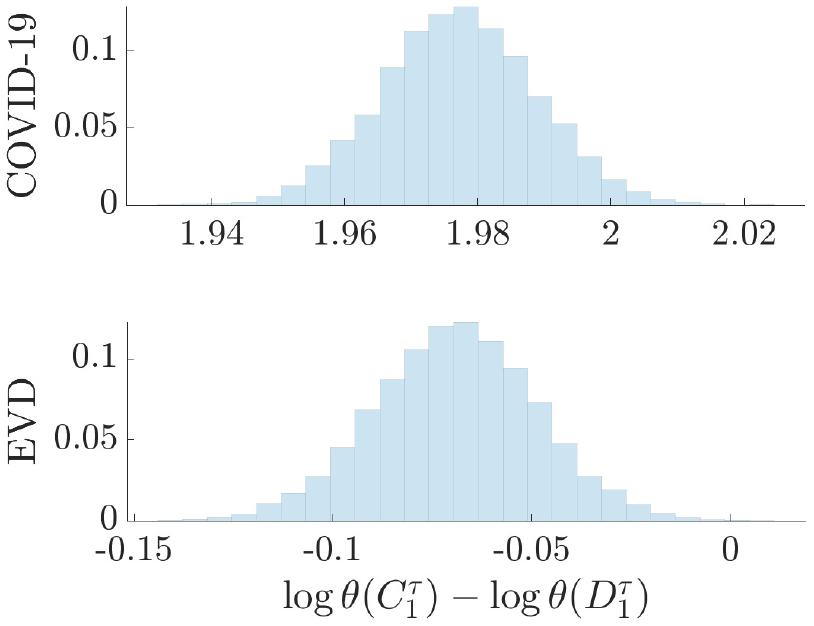
The relative information in case and death data for Ebola virus disease (EVD) and COVID-19 case studies. We compute information metrics *θ*(.) for case 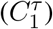 and death 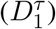 time series using empirically derived under-reporting and delay noise distributions for COVID-19 (top panel) and EVD (bottom panel). See the main text for specific distributions used, which account for the uncertainty in noise estimates (in the absence of knowledge of this uncertainty maximum entropy distributions are applied). We take 10^4^ samples from each distribution and compute the logarithmic difference in the values of our metrics. We find that death data are likely less reliable for COVID-19 (positive values) but more reliable for EVD (negative values).

While we tried to keep estimates as realistic as possible the point of Fig. 5 is to demonstrate how our metrics may be practically applied given noise estimates. Sampling from appropriate distributions means we can propagate the uncertainty on those estimates into our metrics. We provide open source code for modifying this template analysis to include any user-defined distributions in https://github.com/kpzoo/information-in-epidemic-curves. As high-resolution outbreak data collection initiatives such as global.health [50] and REACT [7] progress, enhancing surveillance and our quantification of noise sources, we expect our framework to grow in practical utility.

## Discussion

Public health policymaking is becoming progressively data-driven. Key infectious disease parameters [6] such as instantaneous reproduction numbers and growth rates, fitted to heterogeneous outbreak data sources (e.g., case, death and hospitalisation incidence curves), are increasingly contributing to the evidence base for understanding pathogen spread, projecting epidemic burden and designing effective interventions [4, 6, 51]. However, the validity and value of such parameters depends substantially on the quality of the available surveillance data [1, 7]. Although many studies have made important advances in underscoring and correcting errors in these data [12, 30] no research (to our knowledge) has yet aimed to directly and generally quantify epidemic data quality.

Here we have made some progress towards this aim. We applied Fisher information and experimental design principles to derive a novel framework for quantifying the information within common outbreak data when inferring pathogen transmissibility. Our approach involved finding the total information, 𝕋(.), available from epidemic curves corrupted by reporting delays and under-reporting, which are predominant noise sources that limit surveillance quality. By maximising 𝕋(.), we minimise the overall uncertainty of our transmissibility estimates, hence measuring the reliability of that data stream.

This approach yielded a new non-dimensional metric *θ*(.) that allows analytic and generalisable insights into how noisy surveillance data degrades estimate precision. Using this metric we characterised the impact of different types of delay and under-reporting schemes. We demonstrated that under mean surveillance constraints, constant under-reporting of cases minimises loss of information. However, constant delays in reporting maximise this loss. The first result bolsters conventional thinking [9], while the second highlights the need for timely data [40].

Importantly, our metric provided insight into the nuances of noise, elucidating how the mean and variability of schemes both matter. For example, fluctuating reporting protocols with larger mean may outperform more stable ones at lower mean. Exploiting this and the flexibility of our framework, which can describe the noise in cases, death counts, hospitalisations, infection prevalence and wastewater virus surveys, we demonstrated how diverse data sources might be ranked. Specifically, we critiqued a common assertion about death and case data.

Because the reporting of cases can vary significantly when tracking acute diseases such as COVID-19, various studies have assumed death data to be more reliable [1]. Using our metrics, we presented one of the first qualifications of this claim. We found that the infection fatality ratio (ifr) acts as a reporting fraction with very small mean. Only the most severely varying case reporting protocols can cause larger information loss, suggesting that in many instances this assertion may not hold. Note that this analysis does not even consider the additional advantages that case data bring in terms of timeliness.

However, there may be other crucial reasons for preferring to estimate pathogen spread from death data. For example, if extremely little is known about the level of reporting (very limited surveillance capacity might cause insurmountable case reporting fraction uncertainties) or if a death-based reproduction number is itself of specific interest as a severity indicator [20]. Our framework can also help inform these discussions by improving the precision of our reasoning about noise. This is exemplified by our EVD analysis, where we could show that the large ifr of the disease translated into death counts being the better data, provided their under-reporting is not large.

As hospitalisation curves generally interpolate among the types of noise in case and death data, this might be the best *a priori* choice of data for inferring transmissibility. Some studies also propose to circumvent these ranking issues by concurrently analysing multiple data streams [31, 51]. This then opens questions about how each data stream should be weighed in the ensuing estimates. Our framework may also help by quantifying the most informative parts of each contributing stream. A common way of deriving consensus weighs individual estimates by their inverse variance [52]. As the Fisher information defines the best possible inverse variance of estimates, our metrics naturally apply.

While our framework can enhance understanding and quantification of surveillance noise, it has several limitations. First, it depends on renewal model descriptions of epidemics [27]. These models assume homogeneous mixing and that the generation time distribution of the disease is known. While the inclusion of more realistic network-based mixing may not improve transmissibility estimates [53] (and this extra complexity may occlude insights), the generation time assumption may only be ameliorated through the provision of updated, high quality line-list data [35, 50]. However, our relative metrics in Eq. (10)-Eq. (11) and Eq. (13)-Eq. (14) are mostly robust to generation time distribution misspecifications (and even changes) as they do not depend on the total infectiousness (Λ_*t*_) terms (these cancel out).

Further, our analysis is contingent on having estimates of the delays, under-ascertainment rates and other noise sources within data streams. These may be unavailable or themselves highly unreliable. If at least some information on their uncertainties is available we can propagate these into our metrics by replicating the Monte Carlo approach underlying our case studies. If no estimates are available then we cannot perform any analyses as *R*_*t*_ will not be identifiable. However, our framework can still be of use as a rigorous testbed for examining hypotheses on potential noise sources without extensive simulation.

Recent initiatives have aimed at improving the resolution and completeness of outbreak data [7, 50]. Concurrently, estimating noise sources from both existing and novel data streams is a growing research area [18, 54]. As a result, we expect that our metrics will only increase in practical utility and that concerns around the availability of noise estimates will diminish. We also assumed that the time scale *t* chosen ensures that *R*_*t*_ parameters are independent. This may be invalid but in such instances we can append non-diagonal terms to Fisher information matrices or use our metric as an upper bound.

Last, we defined the reliability or informativeness of a data stream in terms of minimising the joint uncertainty of the entire sequence of reproduction numbers 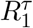. This is known as a D-optimal design [37]. However, we may instead want to minimise the worst uncertainty among the 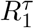 (which may better compensate known asymmetries in inferring transmissibility [55]). Our framework can be reconfigured to tackle such problems by appealing to other design laws. We can solve this specific problem by deriving an *E-optimal* design, which maximises the smallest eigenvalue of our Fisher information matrix.

## Methods

### Renewal models and Fisher information theory

The *renewal model* [27, 36] is a popular approach for describing how infections dynamically propagate during the course of an epidemic. The number of new infections at time *t, I*_*t*_, depends on the instantaneous reproduction number, *R*_*t*_, which counts the new infections generated per infected individual (on average) and the total infectiousness, Λ_*t*_, which measures how many past infections (up to time *t −*1) will effectively produce new ones. This measurement weighs past infections by the generation time distribution, ***w***. We define *w*_*s*_ as the probability that it takes *s* time units for a primary infection to generate a secondary one. The distribution is then 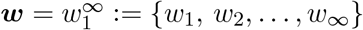.

The statistical relationship between these quantities is commonly modelled as in Eq. (15) with Pois specifying a Poisson distribution [21]. This relationship only strictly holds if *I*_*t*_ is perfectly recorded both in size (no under-reporting) and in time (no delays in reporting).

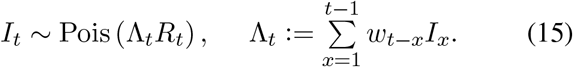

However, as infections are rarely observed, *I*_*t*_ is often approximated by proxies such as reported cases and ***w*** replaced with the serial interval distribution, describing the times between the onset of symptoms of those cases. Eq. (15) has been widely used to model transmission dynamics of many infectious diseases, including COVID-19 [1], influenza [56] and Ebola virus disease [49].

A common and important problem in infectious disease epidemiology is the estimation of the latent variable *R*_*t*_ from the incidence curve of infections or some more easily observed proxy. If this time series persists during 1 ≤*t* ≤*τ*, with *τ* as the present, then we aim to infer the vector of parameters 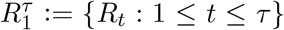 from time series 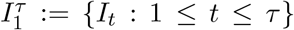 or its proxy (see Results for this more practical inference problem). We assume that time is scaled in units such that *R*_*t*_ can be expected to change (independently) at every *t*. This may be weekly for COVID-19 or malaria [21, 57] but monthly for rabies [58]. Note that ***w*** and *I*_*t*_ must be aggregated, as needed, to match these units. Related branching [59] and moving-average models [60] feature similar aggregation.

Following the development in [44, 56], we solve this inference problem by constructing the incidence log-likelihood function 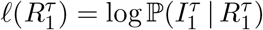 as in Eq. (16) with *K*_*τ*_ as some constant that does not depend on any *R*_*t*_. This involves combining Poisson likelihoods from Eq. (15) across time units 1 ≤ *t* ≤ *τ* as in [21].

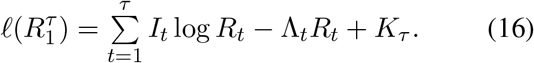

We compute the maximum likelihood estimate (MLE) of *R*_*t*_ as 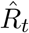, which is the maximal solution of 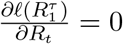. From Eq. (16) this gives 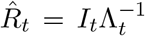 [27]. Repeating this for all *t* we obtain estimates of the complete vector of transmissibility parameters 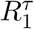 underlying 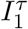.

To quantify the precision (the inverse of the variance, var) around these MLEs or any unbiased estimator of *R*_*t*_ we calculate the Fisher information (FI) that 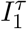 contains about *R*. This is 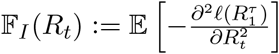, where expectation 𝔼[.] is taken across the data 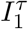 (hence the subscript *I*). The FI defines the best (smallest) possible uncertainty asymptotically achievable by any unbiased estimate, 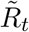. This follows from the Cramer-Rao bound [29], which states that 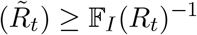. The confidence intervals around var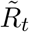 converge to 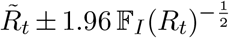. The FI also links to the Shannon mutual information that 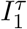 contains about *R*_*t*_ (these measures are bijective under Gaussian approximations) [61, 62] and is pivotal to describing both model identifiability and complexity [29, 34].

Using the Poisson renewal log-likelihood in Eq. (16) we obtain the FI as the left equality in Eq. (17). Observe that this depends on the unknown ‘true’ *R*_*t*_.

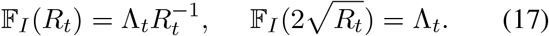

This reflects the heteroscedasticity of Poisson models, where the estimate mean and variance are co-dependent. We construct a square root transform that uncouples this dependence [44], yielding the right formula in Eq. (17). We can evaluate 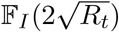 purely from 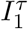. The result follows from the Fisher information change of variables formula 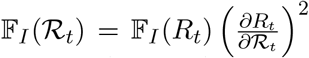 [29]. This transformation has several optimal statistical properties [38, 63] and so we will commonly work with 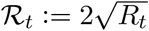.

As we are interested in evaluating the informativeness or reliability of the entire 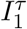 time series for inferring transmission dynamics we require the total FI it provides for all estimable reproduction numbers, 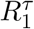. As we noted above, the inverse of the square root of the FI for a single *R*_*t*_ corresponds to an uncertainty (or confidence) interval. Generalising this to multiple dimensions yields an uncertainty ellipsoid with volume inversely proportional to the square root of the determinant of the FI matrix [34, 38]. This matrix has diagonals given by 𝔽_*I*_(*R*_*t*_) and off-diagonals defined as 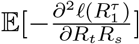 for 1 ≤ *t, s* ≤ *τ*.

Maximising this non-negative determinant, which we denote the total information 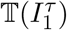 from the data 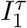, corresponds to what is known as a D-optimal design [37]. This design minimises the overall asymptotic uncertainty around estimates of the vector 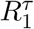. As the renewal model in Eq. (15) treats every *R*_*t*_ as independent, off-diagonal terms are 0 and 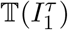 is a product of the diagonal FI terms. Transforming *R*_*t*_ → ℛ_*t*_ we then obtain Eq. (18).

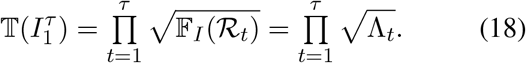

If we work directly in *R*_*t*_ we get 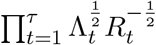 instead. In two dimensions (i.e., *τ* = 2) our ellipsoid becomes an ellipse and Eq. (18) intuitively means that its area is proportional to a product of lengths 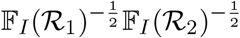, which factors in the uncertainty from each estimate.

We will use this recipe of formulating a log-likelihood for 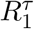 given some data source and then computing the total information, 𝕋(.), it provides about these parameters to quantify the reliability of case, death and other 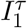 proxies for inferring transmissibility. Comparing data source quality will involve ratios of these total information terms. Metrics such as Eq. (18) are valuable because they measure the usable information within a time series and also delimit the possible distributions that a model can describe given that data (see [34, 64] for more on these ideas, which emerge from information geometry). Transforms like 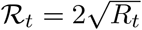 stabilise these metrics (i.e., maximise robustness) to unknown true values [38, 63].

### Epidemic noise sources and surveillance models

We investigate two important and common sources of noise, under-reporting and reporting delay, which limit our ability to precisely monitor 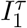, the true time series of new infections. We quantify how much information is lost due to these noise processes by examining how these imperfections degrade 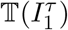, the total information obtainable from 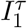 under perfect (noiseless) surveillance for estimating parameter vector 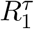 (see Eq. (18)). Fig. 1 illustrates how these two main noise sources individually alter the shape and size of incidence curves.

i. Under-reporting or under-ascertainment. Practical surveillance systems generally detect some fraction of the true number of infections occurring at any given time *t*. If this proportion is *ρ*_*t*_ ≤ 1 then the number of cases, *C*_*t*_, observed is generally modelled as *C*_*t*_ ∼ Bin (*I*_*t*_, *ρ*_*t*_) [23, 57], where Bin indicates the binomial distribution. The under-reported fraction is 1 −*ρ*_*t*_ and so the reported case count *C*_*t*_ ∼ Pois (*ρ*_*t*_Λ_*t*_*R*_*t*_). Reporting protocols are defined by choices of *ρ*_*t*_. Constant reporting (CONR) is the simplest and most popular, assuming every *ρ*_*t*_ = *ρ* [21]. Variable reporting (VARR) describes general time-varying protocols where every *ρ*_*t*_ can differ [28].
ii. Reporting delays or latencies. There can be notable lags between an infection and when it is reported [13]. If ***δ*** defines the distribution of these lags with *δ*_*x*_ as the probability of a delay of *x* ≥ 0 time units, then the new cases reported at *t, C*_*t*_, sums infections actually occurring at *t* but not delayed and those from previous days that were delayed [10]. This is commonly mod-elled as 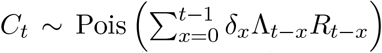 [20, 28] and means that true incidence *I*_*t*_ splits over future times as ∼ Mult(*I*_*t*_, ***δ***), where Mult denotes multinomial [12]. The *C*_*t*_ time series is observed but not yet reported (OBNR) if we later learn about the past *I*_*t*_ splits (right-censoring), else we say data are never reported (NEVR).

We make some standard assumptions [8, 11, 21, 28] in incorporating the above noise sources within renewal model frameworks. We only consider stationary delay distributions i.e., ***δ*** and any related distributions do not vary with time, and we neglect co-dependencies between reporting and transmissibility. Additionally, we assume these distributions and all reporting or ascertainment fractions i.e., *ρ*_*t*_ and related parameters, are inferred from other data (e.g., contact tracing studies or line-lists) [12]. In the absence of these assumptions 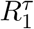 would be non-identifiable and the inference problem ill-defined. In the Results we examine how (i)-(ii) in combination limit the information available about epidemic transmissibility.

## Data Availability

All data and source code (Matlab v2021a) for reproducing the analyses and figures in this manuscript, as well as for applying the methodology we have developed here are freely available at https://github.com/kpzoo/ information-in-epidemic-curves. We include a template function (in Matlab and R) that can be easily modified to compute our metrics with user-defined noise estimates.

https://github.com/kpzoo/information-in-epidemic-curves

## Code Availability

All data and source code (Matlab v2021a) for reproducing the analyses and figures in this manuscript, as well as for applying the methodology we have developed here are freely available at https://github.com/kpzoo/information-in-epidemic-curves. We include a template function (in Matlab and R) that can be easily modified to compute our metrics with user-defined noise estimates.

## Acknowledgments

Thanks to Matthew Hickman for providing useful and interesting comments on the manuscript.

## Author Contributions

Conceptualization, investigation, methodology, formal analysis, funding acquisition and writing (original draft preparation): KVP. Validation: KVP and AEZ. Writing (review and editing): all authors.

## Appendix

### Heterogeneous transmission noise models

For some infectious outbreaks the case (Eq. (1)) and death (Eq. (2)) count time series might be overdispersed [20] i.e., the mean-variance equality inherently assumed by the Poisson renewal model may not be valid. This can result when transmission is strongly influenced by heterogeneities in contacts and infectiousness or when incidence data are corrupted by additional intrinsic noise. These effects are often modelled by generalising Eq. (15) to a negative binomial (NB) form [2, 65], with dispersion *k* and success probability *p*. This leads to the expression below, with mean Λ_*t*_*R*_*t*_ and second argument as *p*.

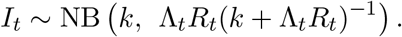

As we focus on the impact of the heterogeneity here, we assume perfect reporting (reporting noise as in the main text only affects the mean of this model). Taking derivatives of the log-likelihood corresponding to the NB model, *ℓ*(*R*_*t*_), we get 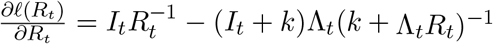. This gives the MLE 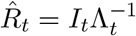, which is the same as for Eq. (15). Computing 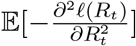 yields the FI that *I*_*t*_ contains about *R*_*t*_ below.

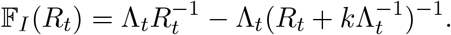

The first term above is the FI from Eq. (17). Heterogeneity therefore subtracts from its FI with a dispersion controlled term. As *k* → ∞ this disappears and NB → Pois. While we do not explicitly include heterogeneity in the analyses of the main text, we do show, importantly, that our results do not qualitatively change if overdispersion is included. There we rank epidemic and death curves, which have been corrupted by under-reporting and delays, according to their Poisson FI. Curves with larger FI are deemed more reliable sources of information about *R*_*t*_. If the delay and under-reporting noise do not alter *k* (i.e., the level of transmission heterogeneity is stable), then this Poisson FI ordering remains valid. We confirm this in Fig. 6, demonstrating a monotonic relationship between the FI from both models at fixed *k*. Our main results are therefore moderately robust to heterogeneities.

**Fig. 6:**
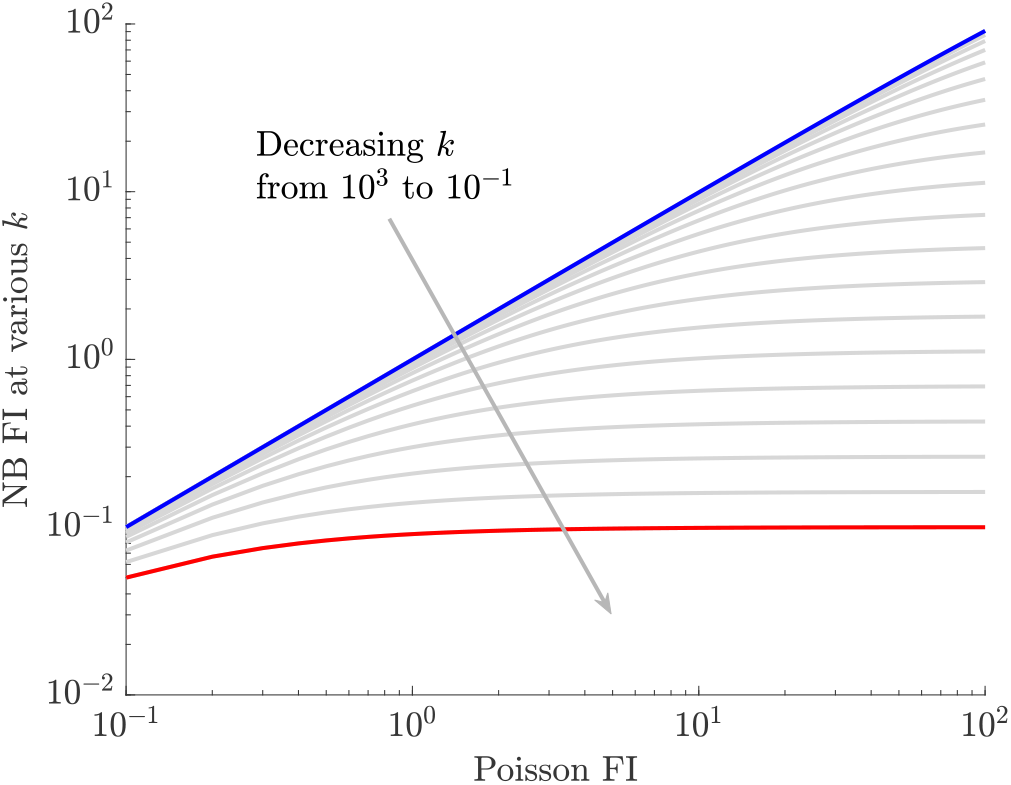
Overdispersion maintains Fisher information based rankings. We plot the FI under a NB observation model that models overdispersion or heterogeneities in transmission, against the FI from the corresponding Poisson renewal model (with the same mean) at *R*_*t*_ = 1. For various dispersion parameters, *k* (smaller *k* means more heterogeneity), we find a monotonic ordering between these two FI values. This suggests that the rankings of data sources derived from their Poisson FI will likely also remain preserved under these more complex NB models, provided those data sources have similar *k* values.

### Information if case source times are never reported

Eq. (5) allows us to compute the FI of under-reported and delayed surveillance data. However, it assumes that we eventually uncover the true time of infection of the delayed cases. This is only valid for OBNR data [12], for which we can separate *C*_*t*_ from Eq. (1) into components 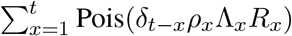 for every *t*. Here we compute the FI for the more complex case of NEVR delays, in which the source data of delayed cases are never reported and hence this sum cannot be decomposed. Specifically, we solve Eq. (6) for a three-dimensional example i.e., *τ* = 3 and prove that we obtain a triangular FI matrix that has diagonal terms as in Eq. (7).

We then compute the total information, 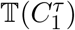 for never reported delays (the NEVR analogue to Eq. (9)) showing that it involves a product of those diagonal terms. This allows us to assert Eq. (9) as an upper bound and to prove convergence of these when the effective reproduction numbers are constant i.e., *R*_*t*_ = *R* for all *t*. The algorithms we apply serve for any *τ* by inductively repeating the procedure here (for code see https://github.com/kpzoo/information-in-epidemic-curves).

We start from Eq. (6) with *α*_(*t*−*x*)*x*_ := *δ*_*t*−*x*_*ρ*_*x*_Λ_*x*_ to define the complete log-likelihood below.

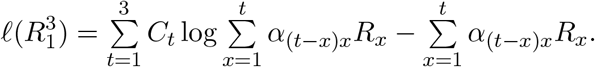

Taking derivatives for *R*_1_ gives 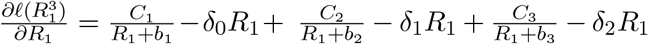 with 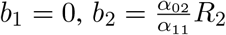 and 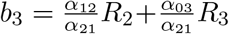. This is repeated for all other *R*_*t*_ and conforms to the description in the main text. When solved and rearranged this gives the MLEs in Eq. (8).

We then calculate 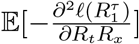 for every combination of *t* and *x* with the expectation of any *C*_*t*_ following as the mean in the Poisson model Eq. (1). This generates the complete FI matrix, 𝔽_*C*_, below with *β*_1_ = *α*_01_*R*_1_, *β*_2_ = *α*_02_*R*_2_ +*α*_11_*R*_1_ and *β*_3_ = *α*_03_*R*_3_ +*α*_12_*R*_2_ +*α*_21_*R*_1_.

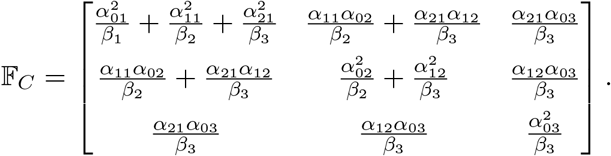

The total information that 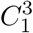 contains about 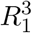 is already computable from the determinant of this matrix. However, we can obtain a more illuminating form by applying elementary column operations.

Such operations do not change the determinant. Using col(*x*) for column *x* we successively apply 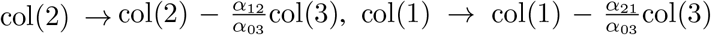 and 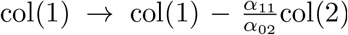, revealing the triangular matrix below. This procedure of subtracting multiples of the later columns from earlier ones can be repeated for any *τ* to also extract analogous triangular forms.

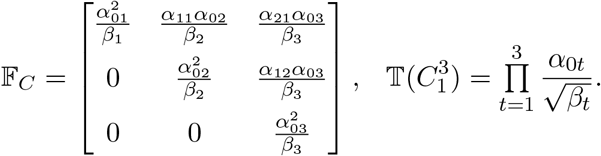

The total information 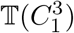 therefore depends on the diagonals of this triangular matrix as shown above.

When written out this corresponds to a product of the terms given in Eq. (7) for all *t*. Each term is smaller than or equal to the corresponding term from an OBNR delay, given in Eq. (5). We can therefore use the total information in Eq. (9) to upper bound that available from a time series with NEVR delays. Intriguingly, this bound is sharp in some important instances. Specifically, if transmissibility is constant so *R*_*t*_ = *R* for all *t* then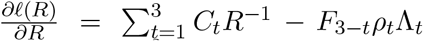 and consequently 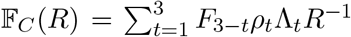. This is precisely the FI obtained under an OBNR delay (by summing Eq. (5)). The MLEs for both delay types are also equal.

